# Extreme COVID-19 waves reveal hyperexponential growth and finite-time singularity

**DOI:** 10.1101/2021.10.15.21265037

**Authors:** Induja Pavithran, R. I. Sujith

## Abstract

Coronavirus disease 2019 (COVID-19) has rapidly spread throughout our planet, bringing human lives to a standstill. Understanding the early transmission dynamics helps plan intervention strategies such as lockdowns that mitigate further spread, minimizing the adverse impact on humanity and the economy^1–3^. Exponential growth of infections was thought to be the defining feature of an epidemic in its initial growth phase^4–7^; any variation from an exponential growth was described by adjusting the parameters of the exponential model^7,8^. Here, we show that, contrary to common belief, early stages of extreme COVID-19 waves display an unbounded growth and finite-time singularity accompanying a *hyperexponential* power-law. The faster than exponential growth phase is hazardous and would entail stricter regulations. Such a power-law description allows us to characterize COVID-19 waves with single power-law exponents, better than piece-wise exponentials. Furthermore, we identify the presence of log-periodic patterns decorating the power-law growth. These log-periodic oscillations may enable better prediction of the finite-time singularity. We anticipate that our findings of hyperexponential growth and log-periodicity will help model the COVID-19 transmission more accurately.

---

Severe acute respiratory syndrome coronavirus 2 (SARS-CoV-2) pandemic, better known as COVID-19, has spread rapidly throughout the globe, since the beginning of January 2020. At the time of writing, it continues to adversely affect millions of people worldwide and has significantly hampered the global economy. Several measures such as closure of international borders and flight bans have been implemented to curtail its transmission. Mitigating the spread of the disease through non-pharmaceutical interventions such as the use of masks, hand sanitizer and the implementation of lockdowns has been critically important^1–3,9^. Imposing appropriate quarantine rules and lockdown measures can reduce the spread of the virus with minimal damage to the economy^10^. Forecasting the dynamics of transmission of the virus can help policymakers to implement appropriate timely interventions^11^.

Epidemiological models have been useful during pandemics and can predict the pattern of transmission of the virus, the number of infections, and the consequences for the near future^12^. The most commonly used model is a susceptible-exposed-infectious-removed (SEIR) model^13^. These models incorporate assumptions from key physical mechanisms involved in the transmission dynamics, the population at risk and the reproductive number of the virus. Besides, a large volume of literature deals with forecasting the pandemic spread using machine learning techniques^14–16^. For modelling and data-driven predictions, one has to rely on the available data and make meaningful inferences.

The standard model for growth of biological populations is the Malthus’ exponential growth model^17^. Most of the studies consider that the number of new infections of COVID-19 increases in an exponential manner^4–6^, except Komarova et al.^16^ who suggested that although the initial phase of the spread is exponential, the long term dynamics may deviate and follow a power-law. A faster than exponential growth phase has been detected during the second wave of COVID-19 in Italy^8^. The accelerated growth was attributed to the limited testing. In this study, we explore all such growth possibilities and discover that the spread of COVID-19 need not necessarily be exponential; it can be faster or slower than exponential from the start of a wave, as observed in several regions for the different waves encountered so far.

Here, we examine data sets of daily infections of COVID-19 for several countries and some of the most densely populated cities. We first illustrate that the exponential model is inadequate to describe the growth of COVID-19 cases. We perform the tests of exponential growth on the data of the 7-day moving average of daily infections (calculated using a sliding window with an overlap of 6 days), which we refer to as *I*. Figure 1 shows the semi-log plot of the 7-day average of the daily infections (in logarithmic scale) as a function of time (linear). An exponential growth rate would reveal a linear increase. In contrast, for some of the waves in the USA, India, Japan and Italy (shown in Fig. 1A), the variation of daily cases depicts a concave upward nature, a characteristic signature of hyperexponential growth^19^. The hyperexponential growth in the transmission dynamics is perhaps not surprising because one encounters hyperexponential growth when dealing with population dynamics, for instance, the growth of human population^19,20^. Further, exponential (Fig. 1B) and slower than exponential (Fig. 1C) growth of daily infections are also observed for different waves of COVID-19 in different countries. The daily infections during the first waves in the UK, Brazil, India and Mexico display a concave downward curve, indicating a slower than exponential growth (Fig. 1C). It is thus evident that the exponential model is not an all encompassing description of the spread of COVID-19.

**Figure 1:**
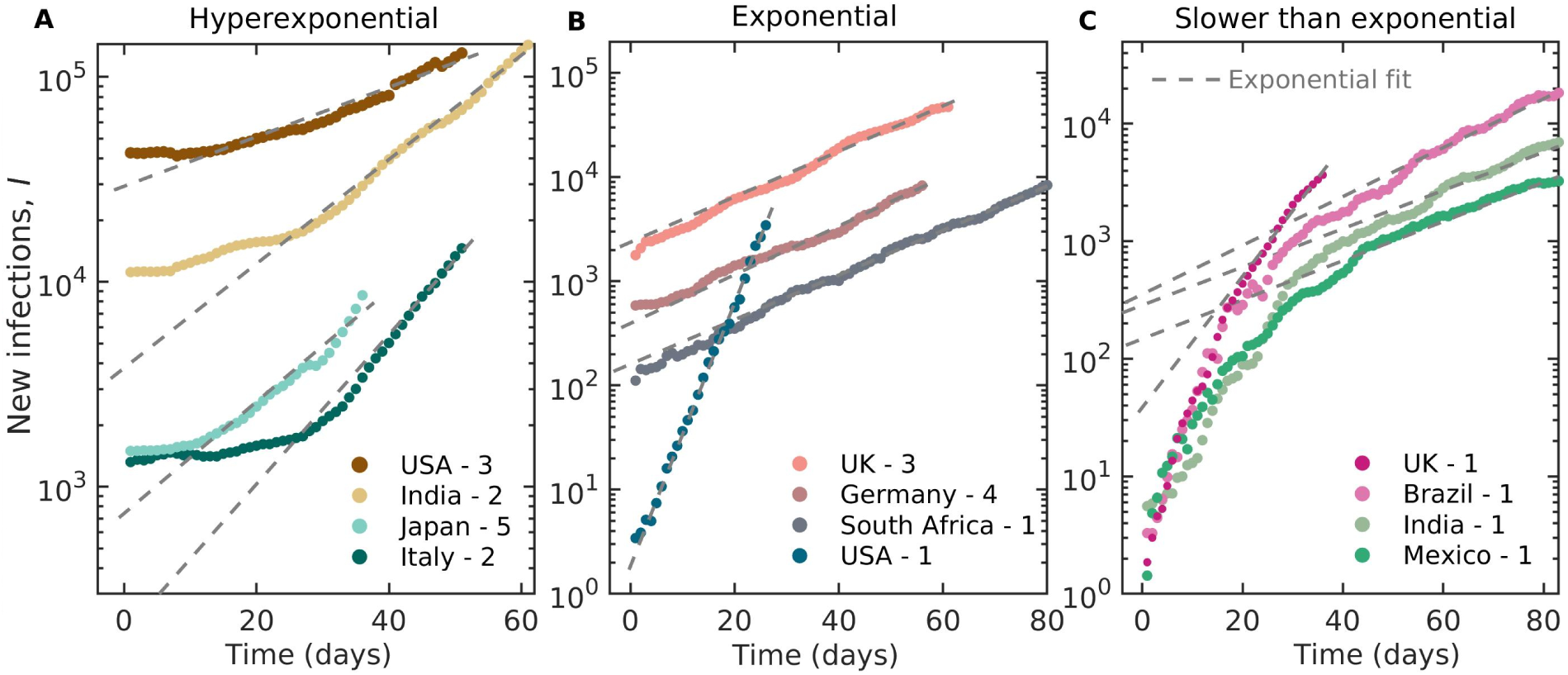
Hyperexponential, exponential and slower than exponential growth of COVID-19 infections. The number of new infections as a function of time in a semi-logarithmic plot reveals three different types of growth behaviour. An exponential growth should appear linear in a semi-log plot. Hyperexponential (i.e., faster than exponential) growth appears as a nonlinear curve which is concave upward (**A**). Such hyperexponential growth is observed for the third wave in the USA (denoted as USA -3 in the figure), India -wave 2, Japan - wave 5 and Italy - wave 2. We show some examples for exponential growth in **B**. The first waves in the UK, India, Brazil and Mexico exhibit concave downward behaviour indicative of slower than exponential growth (**C**). The dotted lines in each plot represent the exponential fit and we can clearly see that the exponential function does not always fit the data. The data is downloaded from^18^. The time (days) are counted from the start of each of the waves and the actual dates corresponding to the data shown are listed in Table. 1 in the methods.

The exponential growth model assumes that the instantaneous rate of change of *I* with respect to time is proportional to *I* itself, i.e., *dI*/*dt* ∝ *I*. We present a more general growth model that can describe faster and slower than exponential growth, as well as exponential growth:

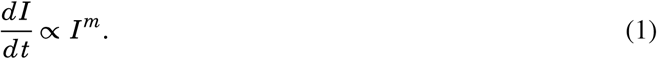

The solution to this equation can exhibit three distinct dynamics depending on the value of *m*.

1. For *m* > 1, *I* = *C*_1_(*t*_*c*_ − *t*)^*α*^ with *α* ∈ (−∞, 0), and *I* → ∞ for *t* → *t*_*c*_ (faster than exponential also known as hyperexponential)^21^.
2. For *m* = 1, 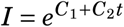(exponential).
3. For *m* < 1, *I* = *C*_1_ + *C*_2_(*t*_*c*_ − *t*)^*α*^ with *α* ∈ (0, ∞), and *I* → *C*_1_ for *t* → *t*_*c*_ (slower than exponential).

Here, *α* and *m* are related as *α* = −1/(*m*−1) and *C*_1_ & *C*_2_ are empirical constants. The critical time *t*_*c*_ is defined as the time at which *I* or any of its derivatives reach infinity. Exponential function is just one specific solution of Eq. 1. We report the presence of all these three types of functions in the data for different COVID-19 waves in various regions. Even though the plots of these functions look similar, careful examination reveals that they are fundamentally different. Solution 2 is an exponential function with a constant exponent, whereas solution 1 is a power-law with faster than exponential growth leading to a finite time singularity at *t*_*c*_.

The singularity originates from the fact that the relative growth rate, (1/*I*) *dI*/*dt*, increases as *I* grows in time for *m* > 1. The relative growth rate of *I*,

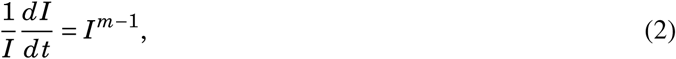

increases as a power-law of *I*, and the doubling-time decreases rapidly when *m* > 1. The successive doubling-time intervals approaches zero rapidly. An infinite number of doubling occurs in a finite time leading to a finite-time singularity. Here, the growth rate (relative to *I*) increases with the number of daily infections for *m* > 1 (Eq. 2). Such a positive feedback asserts that the higher the growth rate of *I*, the higher will be the acceleration of growth (i.e., higher growth of growth rate). Positive feedbacks, when unchecked, can cause runaways until the limiting effects gets triggered. This positive feedback is the basic factor causing a finite-time singularity^21,22^.

For the waves that have hyperexponential growth (the ones shown in Fig. 1A), we fit solution 1. The direct fitting of this equation by minimizing the least square error is highly degenerate, with many solutions which differ by a few percentages in the goodness of fit^19^. Therefore, following Johansen & Sornette^19^, we use a non-parametric approach where we plot log(*I*) as a function of log(*t*_*c*_ − *t*). Such a plot makes evident a linear behaviour for an appropriate *t*_*c*_, which qualifies the power-law as described in solution 1. The slope of this linear curve gives the exponent *α* (see Fig. 2).

**Figure 2:**
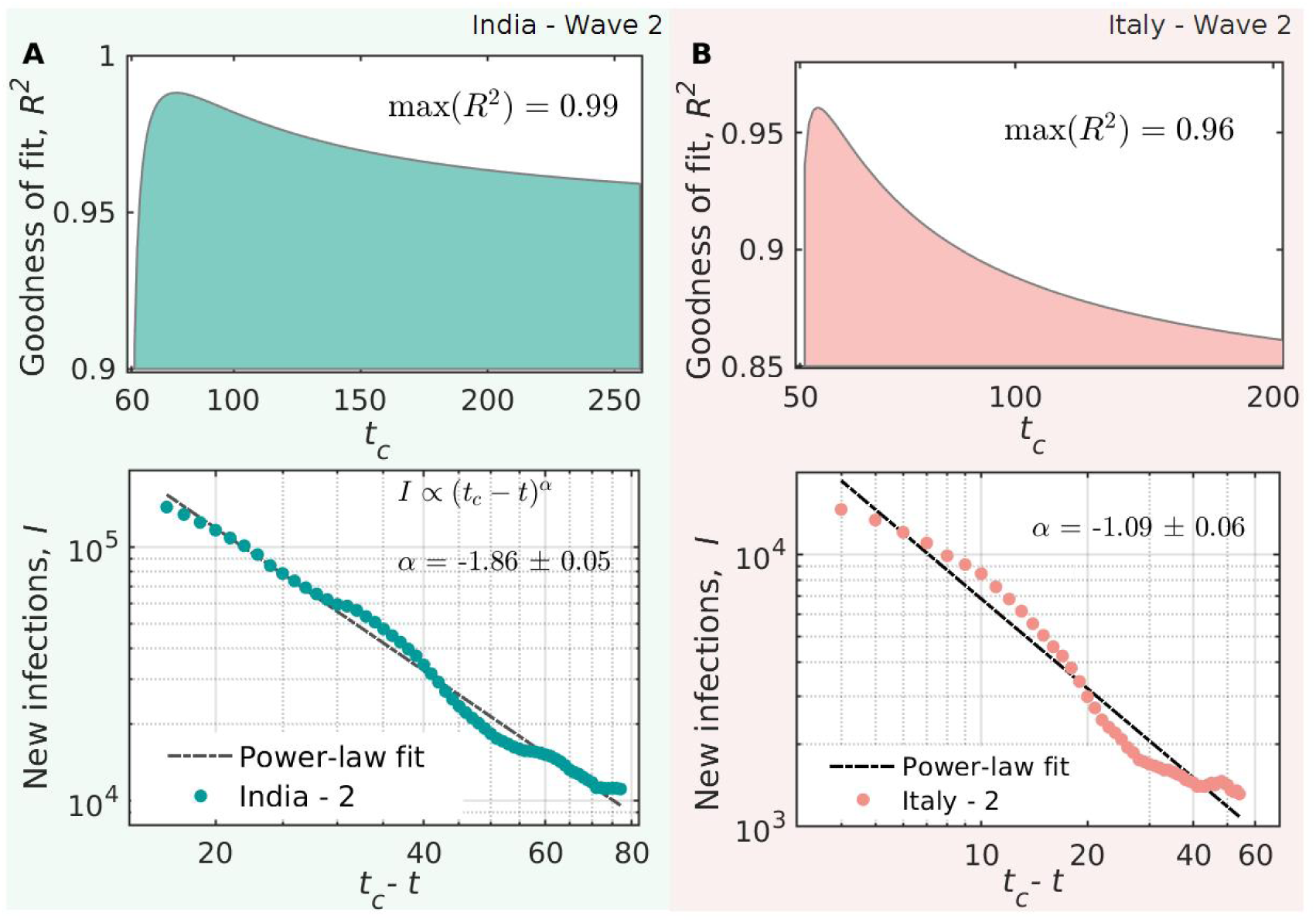
Estimating the critical time *t*_*c*_ of the transmission and the hyperexponential power-law exponent. (**A**) The variation of *R*^2^ of the power-law as a function of the assumed *t*_*c*_ values has a clear peak. We show the the best power-law fit for *I* and *t*_*c*_ − *t* using the *t*_*c*_ estimated by maximizing the *R*^2^ for the second wave in India. (**B**) The second wave of Italy also shows a power-law for the appropriate *t*_*c*_ selected by maximizing the *R*^2^ value. Here, the second wave in Italy has a relatively lower value of *α* compared to India. The *α* indicates the rapidity of the hyperexponential outbreaks.

This fitting procedure is unencumbered by the previously discussed degeneracy and provides a unique and reliable result. We vary *t*_*c*_ over a wide range and calculate the goodness of fit (*R*^2^ values) for all the values of *t*_*c*_ in that range and estimate the max(*R*^2^) to find the best *t*_*c*_ (see Fig. 2). Thus, solution 1 corraborates with the data better than any exponential model. However, *I* cannot rise up to infinity due to multiple reasons. Firstly, the population is not infinite. Then, once the wave begins, there may be lockdowns, isolation of infected people, and other interventions reducing the growth rate of daily infections. Also, those who are infected get immunity, reducing the number of susceptible people. Therefore, *I* deviates from the power-law and the singularity is never reached^23^.

For solution 3, *I* shows a slower than exponential growth, *I* = *C*_1_ + *C*_2_(*t*_*c*_ − *t*)^*α*^ with *α* > 0. Here, ⌈*α*⌉th derivative is a power-law with a finite time singularity at *t*_*c*_ (exponent = (*α* − ⌈*α*⌉) < 0). Therefore, *t*_*c*_ has a physical meaning only for the ⌈*α*⌉th derivative. We restrict ourselves from further investigating such cases to obtain *t*_*c*_ and *α* as they are relatively gradually progressing waves without any finite time singularity in *I* (e.g., the first waves in the UK, Brazil, India and Mexico in Fig. 1C).

In solution 1, the ‘growth rate of daily infections’ (*dI*/*dt*) also follows a hyperexponential power-law with an exponent (*α* − 1). Further, we can see large fluctuations in *dI*/*dt* around the power-law behaviour on approaching *t*_*c*_ (see Fig. 2). These fluctuations can be easily seen in the variation of *dI*/*dt* (Fig. 3B), as compared to *I* (Fig. 3A). These variations are genuine and should not disregarded as noise^24^. We try to explain this by generalizing the power-law exponent *α* to a complex exponent *β* + *iω*. The real part of (*t*_*c*_ − *t*)^*β*+*iω*^ is,

**Figure 3:**
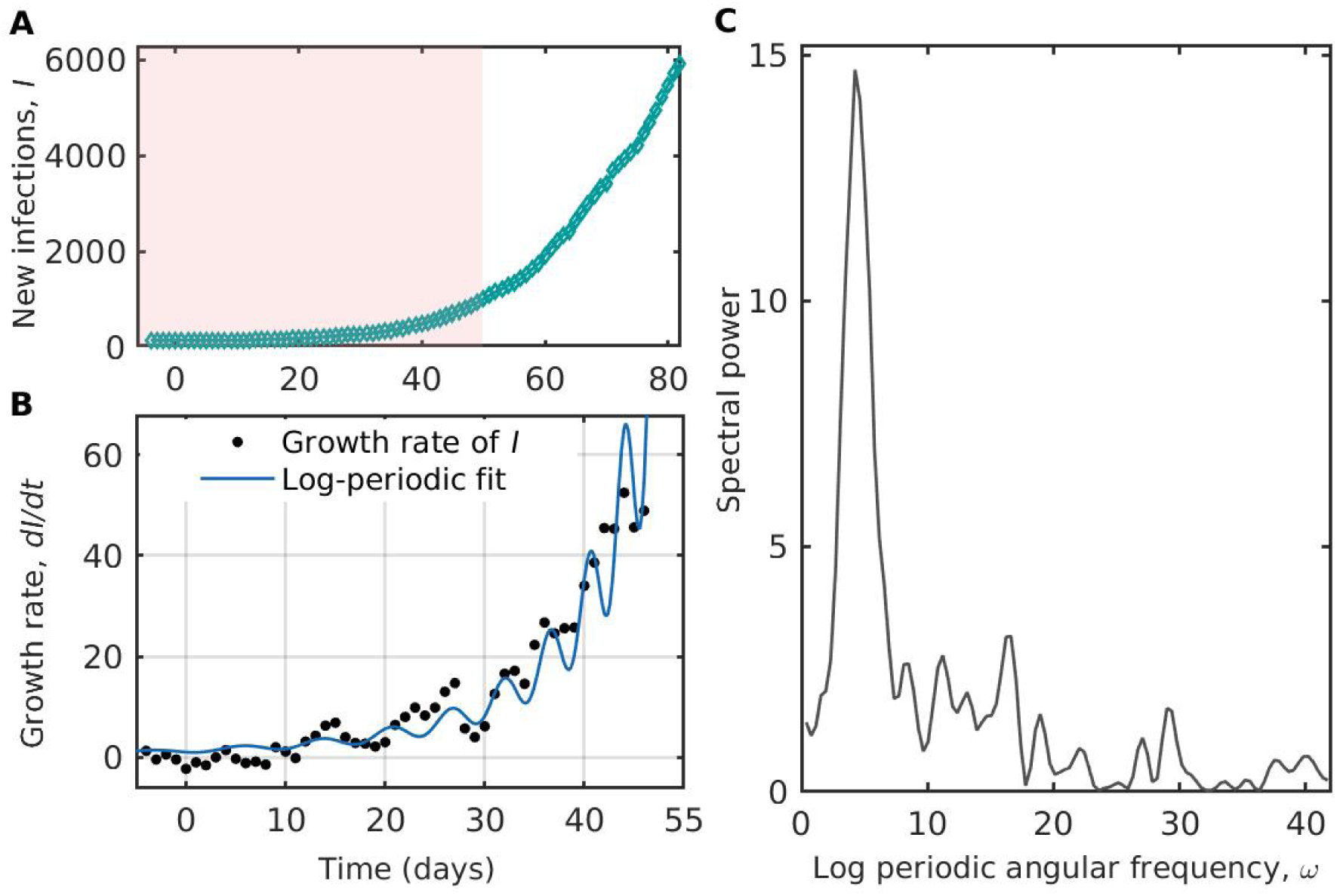
Log-periodicity in the early transmission dynamics of COVID-19. (**A**) We show log-periodicity during the initial hyperexponential growth phase of the second wave in Chennai. The data during the early growth phase shown in the coloured box is used to check log-periodicity. (**B**) We fit the log-periodic power-law to the growth rate of daily infections (*dI*/*dt*) for the city of Chennai in India. We observe log-periodic oscil-lations more clearly in *dI*/*dt*. These fluctuations should not be disregarded as noise. (**C**) A spectral analysis performed on these fluctuations over the power-law confirms that they are not random. Lomb periodogram shows a distinct peak corresponding to *ω* = 4.25. The data for Chennai city is used for this plot^28^.

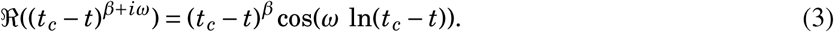

This cosine function decorates the average power-law with oscillations that are periodic in logarithmic scale of (*t*_*c*_ − *t*). These are called *log-periodic* oscillations^24^, and they can explain, to an extent, the observed variability around the power-law. So, considering them may provide better estimates of *t*_*c*_ and *β*. In general, log-periodic oscillations accompany power-laws (both solution 1 and 3) with a critical time *t*_*c*_ where the observable or any of its derivatives tends to infinity in a finite time. These oscillations provide a complementary indication of the impending singularity and are more robust to noise in the data. Note that there can be some extraneous fluctuations in the data, apart from these log-periodic oscillations, caused by some variations in testing or non-pharmaceutical interventions.

There are fundamental rationales behind expecting complex power-law exponents and the associated log-periodic corrections in complex systems^24^. Examples for various mechanisms generating these oscillations include singularities in the Euler equations with complex exponents from the cascade of Rayleigh–Taylor instabilities^25^, speculative bubbles prior to stock market crashes^26,27^, etc. When there are log-periodic structures present in the data, they manifest as oscillations with decreasing time period on approaching *t*_*c*_. The gap between the local maxima of each cycle of oscillations converge to *t*_*c*_ with a geometrical scaling ratio *λ* = exp(2*π*/*ω*). This is related to the existence of a discrete scale invariance with a preferred scaling ratio *λ* as the magnifying factor from one level of the hierarchy to the next^24^.

Log-periodic oscillations are observed in the daily variations of stock market indices. In stock markets, there is a hierarchical structure of imitative behavior among traders. In such a hierarchical structure, a trader influences only a certain number of traders at the same hierarchical level and below^24^. We speculate that the transmission dynamics of infectious diseases such as COVID-19 also follows a hierarchical structure, wherein a person infects a limited number of people, then there are secondary infections from them. Each person in every level has a tree like structure of secondary infections, similar to the top level. For hyperexponential power-laws, ideally, there will be infinite number of infections at a level at *t*_*c*_, when the doubling time is a converging sequence in contrast with the exponential growth.

Next, we will discuss the method to confirm the presence of log-periodicity. Fitting the log-periodic equation with a sufficiently large number of tunable parameters is a delicate problem^24^. Thus, we use a non-parametric test to detect the log-periodic component. The procedure considers only the oscillatory component after removing the accelerating trend. The detrended oscillatory component at any given time *t* is estimated by the difference between the running maximum (using a moving window) till *t* and the running minimum from *t* to the end of the data used^29^. The detrended output, *I*_max_ − *I*_min_, is a non-negative time series; zero corresponds to the position where the maximum value in the past is equal to the minimum value in the future. For instance, if there are no oscillations over the power-law, then *I*_max_ − *I*_min_ will be zero for all *t*. A spectral analysis (Lomb periodogram) is performed on *I*_max_ − *I*_min_ as a function of log(*t*_*c*_ − *t*). Lomb periodogram should have a peak corresponding to the angular frequency *ω* for log-periodic signals. Figure 3C confirms the presence of log-periodicity by detecting a distinct peak, significantly above the noise level, in the Lomb periodogram for the second wave in Chennai. Here, we choose to demonstrate the log-periodicity for a densely populated city, because large countries can have asynchronous appearance of COVID waves in different parts of the country having different policies of interventions. We cannot pinpoint whether the fluctuations are coming from such outside factors or from the underlying discrete scale invariance. However, when we start to see one or two cycles of fluctuations, performing a spectral analysis of the fluctuations can detect the presence of log-periodicity. For example, in the case of the second wave in Chennai, we observe log-periodic oscillations well before the infections grow to very high numbers.

In summary, we detect hyperexponential growth of daily infections in some of the recent COVID-19 waves. We categorize the different growth trends as to whether they are associated with a finite time singularity or not. In most of the countries we have analysed, their severe COVID-19 waves (examples are shown in Methods) show hyperexponential growth with finite time singularity. Singularities are mathematical idealizations of real-world phenomena; they are smoothed out by the finiteness of the system. However, they portend transitions or regime changes. In the context of this pandemic, power-laws with finite time singularities should be interpreted as an indicator of unbounded growth happening over a very short time, similar to phase transitions. Describing pandemic waves with power-laws with a single exponent is more sensible than fitting with an exponential model with varying parameters. Furthermore, we also confirm the presence of log-periodic oscillations in the growth rate of daily infections. This log-periodic correction to the power-law exponent may enable better prediction of *t*_*c*_. Note that hyperexponential behaviour is not a necessary condition for log-periodicity and also hyperexponential power-laws can exist without log-periodicity. Log-periodictity originates from the underlying discrete scale invariance. Finally, we suggest that the mathematical models should not be restricted to describing exponential growth; instead a general growth principle can be considered where the relative growth rate need not be a constant. This would make a better match of models with the real data, thereby enabling better predictions.

## Data Availability

All data produced are available online at https://covid19.who.int and https://data.covid19india.org

https://data.covid19india.org

https://covid19.who.int

## Methods

### Details of the data used

We used the daily infections data for different countries from https://covid19.who.int^1^. We consider the data from the start of a wave; i.e., when the growth rate starts to increase. A seven-day average value of daily infections is used for the analysis throughout this paper,

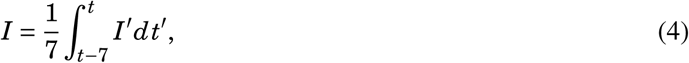

where, *I*′ is the raw tested positive cases in each day.

The details of COVID-19 waves in different countries following hyperexponential, exponential and slower than exponential growth are listed in Table 1. We used the data from the start date to the end date (provided in the Table) of each wave. The corresponding peak daily infections are also provided. Figure 4 shows the number of daily infections as a function of time for six different countries. In all the cases, a hyperexponential growth preceded the peak corresponding to the maximum number of daily infections.

**Table 1:**
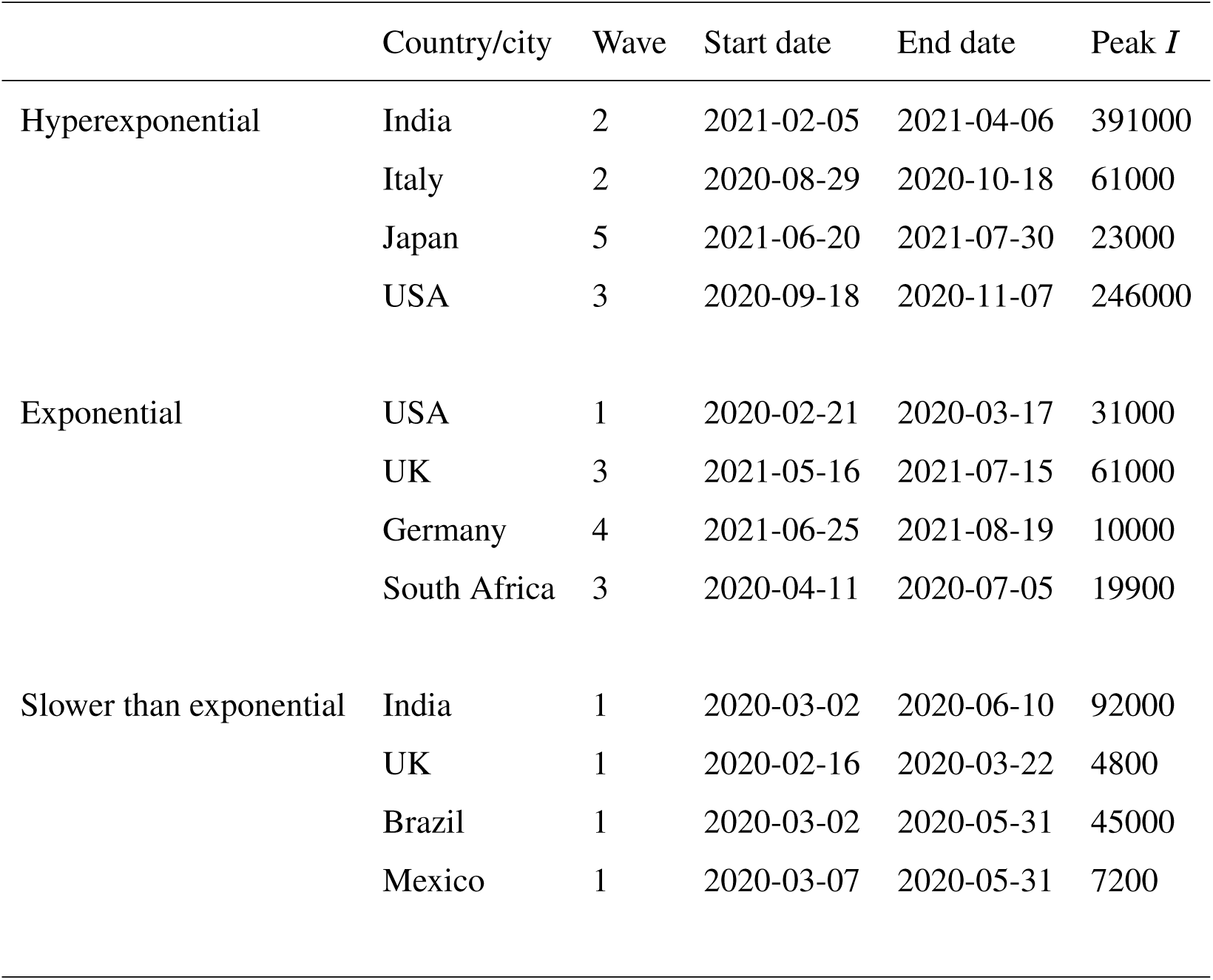
Different waves of COVID-19 considered for the study

| | Country/city | Wave | Start date | End date | Peak $I$ |
| --- | --- | --- | --- | --- | --- |
| Hyperexponential | India | 2 | 2021-02-05 | 2021-04-06 | 391000 |
|  | Italy | 2 | 2020-08-29 | 2020-10-18 | 61000 |
|  | Japan | 5 | 2021-06-20 | 2021-07-30 | 23000 |
|  | USA | 3 | 2020-09-18 | 2020-11-07 | 246000 |
| Exponential | USA | 1 | 2020-02-21 | 2020-03-17 | 31000 |
|  | UK | 3 | 2021-05-16 | 2021-07-15 | 61000 |
|  | Germany | 4 | 2021-06-25 | 2021-08-19 | 10000 |
|  | South Africa | 3 | 2020-04-11 | 2020-07-05 | 19900 |
| Slower than exponential | India | 1 | 2020-03-02 | 2020-06-10 | 92000 |
|  | UK | 1 | 2020-02-16 | 2020-03-22 | 4800 |
|  | Brazil | 1 | 2020-03-02 | 2020-05-31 | 45000 |
|  | Mexico | 1 | 2020-03-07 | 2020-05-31 | 7200 |

**Figure 4:**
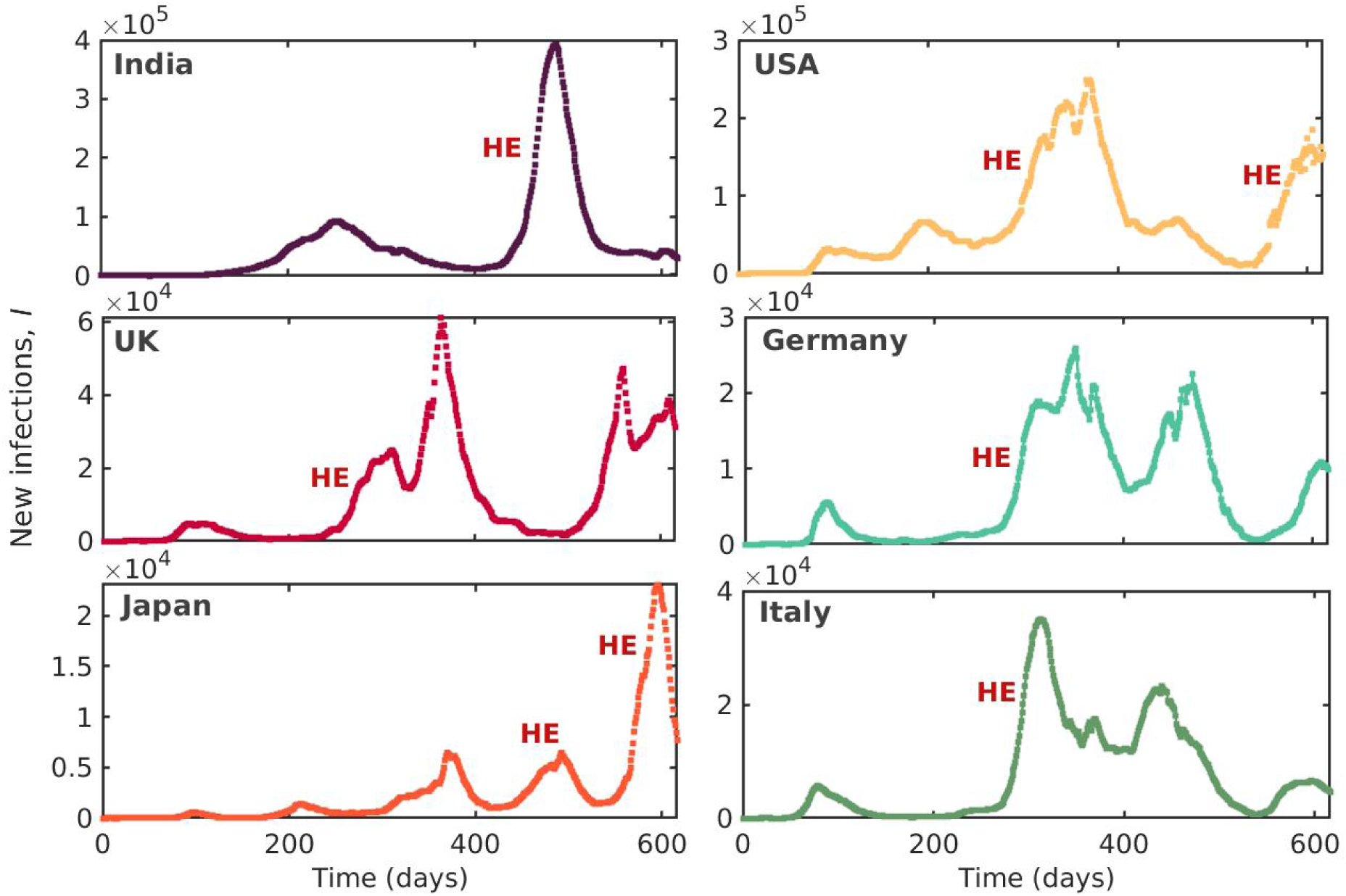
Extreme COVID-19 waves have hyperexponential growth. In each country, their worst COVID-19 waves are hyperexponential in the initial growth phase. Hyperexponential waves are indicated as **HE** in the figure. We show COVID-19 waves of some countries where the highest peak of daily infections is associated with hyperexponential growth. The time (days) for this plot is counted from 03 January 2020 for all the countries.

#### Test for exponential growth

We plot the daily infections in a semi-logarithmic scale (log(*I*) as function of time). A straight line in such a plot represents exponential behaviour. Concave up and concave down plots correspond to hyperexponential and slower than exponential growth of *I*, respectively.

#### Non-parametric approach to estimate *t*_*c*_ for hyperexponential growth

Direct fitting of hyperexponential power-law is difficult as there are several parameters^2^. Thus, following Johansen and Sornette^2^, we plot log(*I*) as a function of log(*t*_*c*_ − *t*) by assuming different values of *t*_*c*_. We find the appropriate *t*_*c*_ for the best fit by maximizing the *R*^2^ value of linear fit for a wide range of *t*_*c*_. The plot of log(*I*) vs log(*t*_*c*_ − *t*) shows a straight line for this *t*_*c*_. The slope of this line gives the exponent *α* (see solution 1 of Eq. 1 in the main text).

## Acknowledgements

We thank all the health care workers around the globe who are working hard to help humanity during this pandemic. We acknowledge our colleagues A. Roy, A. J. Varghese, P. Kasthuri, S. Tandon and A. Banerjee from Indian Institute of Technology Madras for the valuable suggestions on this manuscript.

## Authors contributions

R.I.S. conceived the study. R.I.S. and I.P. conceptualized the study. All authors contributed to the analysis and interpretation of the results and writing of the manuscript.

## Data and materials availability

All the data used are freely available from the websites which are cited appropriately.

## Additional information

